# Thrombotic adverse events reported for Moderna, Pfizer and Oxford-AstraZeneca COVID-19 vaccines: comparison of occurrence and clinical outcomes in the EudraVigilance database

**DOI:** 10.1101/2021.09.12.21263462

**Authors:** Mansour Tobaiqy, Katie MacLure, Hajer Elkout, Derek Stewart

## Abstract

**Background:** Vaccination against COVID-19 is the cornerstone to control and mitigate the ongoing pandemic. Thrombotic adverse events linked to Moderna, Pfizer and the Oxford-AstraZeneca vaccine have been documented and described as extremely rare. While the Oxford-AstraZeneca vaccine has received much of the attention, the other vaccines should not go unchallenged. This study aimed to determine the frequency of reported thrombotic adverse events and clinical outcomes for these three COVID-19 vaccines, namely, Moderna, Pfizer and Oxford-AstraZeneca

**Methods:** A retrospective descriptive analysis was conducted of spontaneous reports for Moderna, Pfizer and Oxford-AstraZeneca COVID-19 vaccines submitted to the EudraVigilance database in the period from 17 February to 14 June 2021.

**Findings:** There were 729,496 adverse events for the three vaccines, of which 3,420 were thrombotic, mainly Oxford-AstraZeneca (n=1,988, 58·1%) followed by Pfizer (n=1,096, 32·0%) and Moderna (n=336, 9·8%). As serious adverse events, there were 705 reports of pulmonary embolism for the three vaccines, of which 130 reports (18·4%) were for Moderna, 226 reports (32·1%) for Pfizer and 349 (49·5%) for Oxford-AstraZeneca vaccines. The occurrence of pulmonary embolism is significantly associated with a fatal outcome (P=<0·001). Sixty-three fatalities were recorded (63/3420, 1.8%), of which Moderna (n=6), Pfizer (n=25) and Oxford-AstraZeneca (n=32).

**Interpretation:** Thrombotic adverse events reported for the three vaccines remains extremely rare with multiple causative factors reported elsewhere as precipitating these events. Practicing vigilance and proper clinical management for the affected vaccines, as well as continuing to report adverse events, are essential.

**Funding:** No funding was sought for this study.

**Research in context:** *Evidence before this study:* During the first quarter of 2021, several European countries suspended the use of the Oxford–AstraZeneca vaccine amid reports of blood clot events and the death of a vaccinated person. This was followed by several reports of fatalities related to pulmonary embolism and other thrombotic events including thrombocytopenia which has been referred to as vaccine-induced immune thrombotic thrombocytopenia (VITT). The European Medicines Agency on 18 March 2021 concluded that the Oxford– AstraZeneca vaccine was safe, effective and the benefits outweighed the risks.

*Added value of this study:* This study investigated the occurrence of thrombotic adverse events and their clinical outcomes of the three approved and most used COVID-19 vaccines namely Moderna, Pfizer and Oxford-AstraZeneca, using one of the largest spontaneous adverse events databases, namely EudraVigilance. Out of 729,496 adverse events reported for the three vaccines in the study period, only 3420 (0.47%) potential thrombotic adverse events were reported, the majority associated with Oxford-AstraZeneca (n=1,988, 58.1%).

*Implications of all the available evidence:* More than 4·89 billion doses of different COVID-19 vaccines have been administered across the globe. Despite thrombotic adverse events reported for the three vaccines in focus for this study - Moderna, Pfizer and Oxford-AstraZeneca - being extremely rare, so continuing to report adverse events is essential. On the basis of scientific evidence showing that benefit outweighs risk, people continue to be urged to accept the vaccination when offered.

## Introduction

Vaccination against COVID-19 is the cornerstone to control and mitigate the ongoing pandemic.^1^ Thrombotic adverse events including venous thrombosis, thrombocytopenia and ischemic stroke which have been reported in patients in the days following their first dose of vaccine with varying consequences.^2-4^ Thrombocytopenia has been reported as being associated with the Oxford-AstraZeneca vaccine.^4^ This is a clinical syndrome which manifests as a high level of antibodies against platelet factor 4 (PF4) unrelated to heparin-induced thrombocytopenia (HIT). It is referred to as vaccine-induced immune thrombotic thrombocytopenia (VITT). Similar to HIT, bleeding is rare with both HIT and VITT, however, both are associated with thromboembolic complications involving the arterial and venous systems.^4,5^ A rare arterial thrombosis may also be considered an adverse event associated with COVID-19 vaccination as reported in a recent case of malignant cerebral infarct that has been associated with thrombocytopenia.^6^ This condition manifests as high serum levels of antibodies to PF4–polyanion complexes which has been observed in two young, healthy, adult women within 10 days of receiving Oxford-AstraZeneca vaccination.^6^ While the Oxford-AstraZeneca vaccine has been the focus, particularly for blood clotting events, other vaccines have also been reported for thrombotic adverse events. This has included Johnson & Johnson’s Janssen (J&J/Janssen) COVID-19 Vaccine, that was suspended by the Food and Drug Administration (FDA) in the United States (US) and then resumed when FDA concluded that its’ benefits outweighed the known and potential risk.^7^ A recent pharmacovigilance study reported 28 potential thrombotic adverse events out of 54,571 patient vaccinations linked to the Oxford-AstraZeneca vaccine giving an occurrence rate of 0·05%. The study was also based on data from the EudraVigilance (EV) database in the period from 17 February to 12 March in 2021.^8^ Among these, three fatalities were related to pulmonary embolism with one fatality to thrombosis. The same study identified 29 deep venous thrombosis (DVTs) associated with Moderna vaccine, of which 12 also had pulmonary embolism; there were no reported deaths or fatal events but 13 DVTs with Pfizer vaccine in the same study period.^8^

Reports comparing occurrence and clinical outcomes of thrombotic adverse events with these three vaccines remain scare. Adverse events related to vaccines are the most frequently reported among the collected spontaneous reports logged with the EV database.^9^ As COVID-19 vaccination is likely to be broadened to younger age groups, with talk of top-up vaccination for older age groups, decision makers need current evidence on which to base vaccine recommendations. Therefore, the aim of the present study was to compare the reported occurrence of thrombotic adverse events and clinical outcomes for three COVID-19 vaccines, namely Moderna, Pfizer and Oxford-AstraZeneca, based on reports from the EudraVigilance database with a particular focus on pulmonary embolism.

## Methods

A retrospective descriptive analysis was conducted of COVID-19 vaccines spontaneous reports for Moderna, Pfizer and Oxford-AstraZeneca COVID-19 vaccines submitted to the EudraVigilance database for the period 17 February to 14 June 2021.

### Data sources and setting

Data were extracted from EV a European Economic Area (EEA) wide open access pharmacovigilance database which collates adverse event reports from non-healthcare and healthcare professionals. The line listing section of spontaneous reports submitted to the EV for COVID-19 vaccines Moderna, Pfizer (Tozinameran) and Oxford-AstraZeneca (ChAdOx1-S) was accessed. The following search terms were applied: thrombosis; embolism; thromboembolism; embolic and, thrombotic. Taking pulmonary embolism as a focus, serious adverse events were identified and further analysed according to age, sex and type of vaccine. The reports identified were then exported to Microsoft Excel files and classified according to the following variables: age (age groups: ≥85 years old, 65-84 and 18-64 years old), sex, month of reporting, origin of the report, reporter’s profession (healthcare professional or non-healthcare professional) and concomitant conditions. Data were tabulated and presented along with the clinical outcomes which were categorised into four sections: (i) recovered, (ii) recovering, (iii) not yet recovered, and (iv) fatal outcome.

Ethical review was not required for the open access data. The access policy of European Medicines Agency (EMA) states that “No authorisation for accessing the ICSR (Level 1) data set by means of the adrreports.eu portal is required i.e. all academic researchers can access adverse reaction data of interest.^9^”

### Statistical analysis

Data were analysed using descriptive statistics to determine the study population characteristics. Variables were reported as absolute numbers and percentages. All analyses were performed using SPSS for Windows 20·0 (SPSS, Chicago, Illinois, USA). Differences in proportions between the groups were compared with the Chi-square test and monthly trends in reporting were analysed. A P-value of less than 0·05 was considered statistically significant.

## Results

### Frequency of adverse events

During the study period, a total of 729,496 adverse events were reported for the three vaccines: Oxford-AstraZeneca (n=337,712; 46·3%), Pfizer (n=311,364; 42·7%) and Moderna (n=80,420; 11·0%).

### Frequency of thrombotic adverse events associated with the three vaccines

As shown in Table 1,of those reports, there were 3,420 thromboembolic adverse events reported, with 1,988 (58.1%) for Oxford-AstraZeneca, 1096 (32%) for Pfizer and 336 (9.8%) for Moderna. Table 1 gives the general characteristics of the reports and Table 2 the number of monthly reports including reports with a fatal outcome.

**Table 1.** Summary of demographics of EudraVigilance database thrombotic adverse event reports for Moderna, Pfizer and Oxford-AstraZeneca vaccines

| Variable |  | n (%) |  |  |
| --- | --- | --- | --- | --- |
|  |  | Moderna<br>336 (9.8) | Pfizer<br>1096 (32) | Oxford-AstraZeneca<br>1988 (58.1) |
| Sex | Male | 171 (50.9) | 494 (45.1) | 952 (47.9) |
|  | Female | 164 (48.8) | 583 (53.2) | 1006 (50.6) |
|  | Not specified | 1 (0.3) | 19 (1.7) | 30 (1.5) |
| Reporter profession | Healthcare professional | 293 (87.2) | 804 (73.4) | 1422 (71.5) |
|  | Non-healthcare professional | 43 (12.8) | 292 (26.6) | 566 (28.5) |
| Geographical area | EU | 96 (28.6) | 768 (70.1) | 877 (44.1) |
|  | Non-EU/EEA* | 240 (71.4) | 328 (29.9) | 1111 (55.9) |
| Age group of patients | 18-64 years | 140 (41.7) | 374 (34.1) | 1069 (53.8) |
|  | 65-84 years | 162 (48.2) | 519 (47.3) | 725 (36.4) |
|  | 85 years and older | 27 (8.0) | 147 (13.4) | 81 (4.1) |
|  | Missing data | 7 (2.1) | 56 (5.1) | 111 (5.6) |

\*EEA – European Economic Area

**Table 2.** Monthly distribution of thromboembolic EV reports for the three vaccines from 17 February to 14 June 2021

|  | Number of reports | Reports with fatal outcome |
| --- | --- | --- |

| Month | Moderna | Pfizer | Oxford-AstraZeneca | Moderna | Pfizer | Oxford-AstraZeneca |
| --- | --- | --- | --- | --- | --- | --- |
| February | 6 | 24 | 0 | 0 | 0 | 0 |
| March | 29 | 127 | 319 | 0 | 0 | 4 (1.3) |
| April | 47 | 285 | 591 | 2 (4.3) | 7 (2.5) | 2 (0.3) |
| May | 169 | 481 | 758 | 4 (2.4) | 8 (1.7) | 15 (2.0) |
| June | 85 | 173 | 319 | 0 | 10 (5.8) | (3.1) |

### Pulmonary embolism reports

There were 705 reports of pulmonary embolism during the study period for the three vaccines that included deep vein thrombosis with and without pulmonary embolism. There were 130 reports (38·7%) for Moderna vaccine, 226 reports (20·6%) for Pfizer and 349 reports (49·5%) for Oxford-AstraZeneca vaccine (Table 3). Occurrence of pulmonary embolism is significantly associated with a fatal outcome (P=<0·001).

**Table 3.** Clinical outcome of adverse events and demographic variables related to Moderna, Pfizer and Oxford-AstraZeneca vaccines with a focus on pulmonary embolism from 17 February to 14 June 2021

|  | Clinical outcome of all adverse event reports<br>n (%) |  |  |  |  | Pulmonary<br>Embolism<br>only<br>n (%) |
| --- | --- | --- | --- | --- | --- | --- |
|  | Outcome<br>unknown | Recovered | Recovering | Not yet<br>recovered | Fatal |  |
| **Type of vaccine** |  |  |  |  |  |  |
| Moderna (N=336) | 96 (28.6) | 50 (14.9) | 40 (11.9) | 144 (42.9) | 6 (1.8) | 130 (38.7) |
| Pfizer (N=1096) | 146 (13.3) | 125 (11.4) | 449 (71.0) | 351 (32.0) | 25 (2.3) | 226 (20.6) |
| Oxford-AstraZeneca (N=1988) | 347 (17.5) | 194 (9.8) | 790 (39.7) | 625 (31.4) | 32 (1.6) | 349 (17.6) |
| **Age group of patients** |  |  |  |  |  |  |
| 18-64 years (N=1583) | 261 (16.5) | 167 (10.5) | 573 (36.2) | 566 (35.8) | 16 (1.0) | 284 (17.9) |
| 65-84 years (N=1406) | 238 (16.9) | 153 (10.9) | 559 (39.8) | 421 (29.9) | 35 (2.5) | 343 (24.4) |
| 85 years and older (N=255) | 48 (18.8) | 30 (11.9) | 82 (32.2) | 87 (34.1) | 8 (3.1) | 56 (22.0) |
| Not specified (N=174) | 42 (24.1) | 18 (10.3) | 65 (37.4) | 45 (1.3) | 4 (2.3) | 22 (12.6) |
| **Sex** |  |  |  |  |  |  |
| Male (N=1617) | 268 (16.6) | 181 (11.2) | 637 (39.4) | 507 (31.4) | 24 (1.5) | 373 (23.1) |
| Female (N=1753) | 313 (17.9) | 183 (10.4) | 615 (35.1) | 603 (34.4) | 39 (2.2) | 323 (18.4) |
| Not specified (N=50) | 8 (16.0) | 4 (8.0) | 28 (56.0) | 10 (20.0) | .. | .. |

### Outcome of the thrombotic adverse events

Of the total 705 reports, 63 reports were for a fatal outcome; of those, 6 cases (1·8%) were for the Moderna vaccine, 25 (2·3%) for Pfizer vaccine and 32 reports (1·6%) for the Oxford-AstraZeneca vaccine. Out of the total 63 reports of fatal outcomes 38 (60·3%) had pulmonary embolism compared to 25 (39·7%) fatal reports for other thrombolytic adverse events. Within the type of vaccine, of the 38 reports with fatal outcome and pulmonary embolism, 3 of 6 reports were recorded for Moderna, 17 out of 25 for Pfizer and 18 out of 32 reports for Oxford-AstraZeneca vaccines comprising 50·0%, 68·0% and 56·3% of the total reports with fatal outcome for each vaccine, respectively.

Table 3 presents the number and proportions of the range of outcomes among the three COVID-19 vaccines. Moreover, the analyses also showed 589 reports with an unknown outcome, with Moderna (n=96; 28·6%), Pfizer (n=146; 13·3%), and for Oxford-AstraZeneca (n=347; 17·5%) vaccines; 369 reports for recovered patients for Moderna (n=50; 14·9%), for Pfizer (n=125; 11·4%) and for Oxford-AstraZeneca (n=194; 9·8%) vaccines; 1279 reports for recovering patients with Moderna (n=40; 11·9%), for Pfizer (n=449; 71%), and for Oxford-AstraZeneca (n=790; 39·7%) vaccines. 1120 reports were defined as having a not recovered outcome with Moderna (n=144; 42·9%), for Pfizer (n=351; 32%) and for Oxford-AstraZeneca (n=625; 31·4%) vaccines.

Regarding age, those in the 65-84 years age group reported higher fatal outcomes than other age groups (55·5% of all deaths). When examined for each type of vaccine, about half of the reports with a fatal outcome occurred in the same age group. However, only Moderna vaccine showed significant association between age and a fatal outcome (P value = 0·008) (Tables 3-4). In addition, the reports contained other concomitant adverse events such as thrombocytopenia, arrhythmia, and cerebro-vascular thrombosis (Table 5); however, data on concomitant drugs was unavailable.

**Table 4.** Summary of reports with fatal outcome in relation to age and sex for each type of vaccine

| Type of vaccine<br>Fatal Outcome | Sex n (%) |  | Age n (%) |  |  |  |
| --- | --- | --- | --- | --- | --- | --- |
|  | Male | Female | Not specified | 18-64 | 65-84 | 85 and over |
| Moderna | 1 (1.6) | 5 (3.0) | .. | .. | 4 (2.5) | 2 (7.4) |
| Pfizer | 10 (2.0) | 15 (2.6) | .. | 5 (1.3) | 17 (3.3) | 3 (2.0) |
| Oxford-AstraZeneca | 13 (1.4) | 19 (1.9) | 4 (3.6) | 11 (1.0) | 14 (1.9) | 3 (3.7) |

**Table 5:** Concomitant clinical conditions and other thrombotic adverse events

| Concomitant | Moderna | Pfizer | Oxford-AstraZeneca | Total |
| --- | --- | --- | --- | --- |
| Thrombocytopenia | 6 (1.8) | 18 (1.6) | 136 (6.8) | 157 |
| Arrhythmia | 7 (2.1) | 5 (0.5) | 8 (0.4) | 29 |
| Cerebro-vascular thrombosis | 4 (1.2) | 9 (0.8) | 12 (0.6) | 20 |

## Discussion

The development of safe, effective, affordable vaccines against COVID-19 remains the cornerstone to mitigating this pandemic. By 20 August 2021, more than 4·89 billion doses of different COVID-19 vaccines have been administered across the globe, and over 40 candidate vaccines were in human trials,^10,11^ and yet there is variable hesitancy, fear and anxiety^12^ of perceived risk of vaccination.^13^ A recent UK study found increased risks of both haematological and vascular events that led to hospital admission or death following first doses of both Pfizer and Oxford-AstraZeneca vaccines but also noted the risk was higher following COVID-19 infection.^14^

This study identified 729, 496 adverse events for the three vaccines under investigation (Moderna, Pfizer and Oxford-AstraZeneca) for the period of study from 17 February to 14 June 2021, of which 3,420 thrombotic adverse events: 336 (0·41%) for Moderna, 1096 (0·35%) Pfizer and 1988 (0·58%) for Oxford-AstraZeneca vaccines. In this study, thrombocytopenia with concomitant clinical conditions were reported 157 times with the three vaccines and was associated mostly with Oxford-AstraZeneca (n=134, 85·3%) than with Pfizer (n=17, 10·8%) and Moderna (n=6, 3·8%). This finding has been demonstrated recently from a study that reported thrombotic events in 11 patients post-vaccination with Oxford-AstraZeneca and showed evidence of immune thrombotic thrombocytopenia mediated by high level of platelet-activating antibodies against PF4.^3^ In this study, the majority (53·8%) of potential thrombotic adverse events that were linked to Oxford-AstraZeneca vaccine were reported in the age group 18-64 years, whereas the majority of these events were reported in the age group of 65-84 years for Moderna (48·2%) and Pfizer vaccine (47·3%) respectively. In contrast, other research which used EV database, showed that reported thrombotic events were more prevalent in the age group <65 among Oxford-AstraZeneca vaccine’s recipients than Moderna or Pfizer vaccine recipients.^15^

Of note, there was a slight increase, not statistically significant, in the number of reports of female sex for Oxford-AstraZenaca (50·6%) and Pfizer vaccine (53·2%). However, the Moderna vaccine had more reports linked to males (P-value 0.008). We have reported earlier approximately double the occurrence of potential thrombotic events reported in females (n=19) than males (n=9) for Oxford-AstraZenaca vaccine.^8^ Similarly, the clinical and laboratory data of 11 thrombotic patients post vaccination with Oxford-AstraZeneca were 9 female patients with a median age of 36 years (range 22 to 49).^16^

Most potential thrombotic adverse events were reported for Oxford-AstraZeneca (n=1,988; 58.1%). However, the present study and other pharmacovigilance research demonstrated that thrombosis with thrombocytopenia appears to occur with all three vaccines, with higher rates in those who had received the Oxford-AstraZeneca vaccine.^15,17^ It has been suggested that administration of Oxford-AstraZeneca vaccine should be considered carefully and only when the potential benefit outweighs any potential risks, particularly in patients with a history of cerebral venous sinus thrombosis, acquired or hereditary thrombophilia, heparin-induced thrombocytopenia.^18,19^

Pulmonary embolism (PE) is an acute complication of DVT and is consider a serious adverse event,^20^ and the third most common cause of cardiovascular death.^21^ In this study, among the severe clinical outcomes associated with the three vaccines, 705 reports included pulmonary embolism with 63 fatalities; however, this finding should be interpreted with great caution given the nature of the present data and the passive reporting system used.

The limitations of this study, as with spontaneous reporting schemes of adverse reactions of both drugs and vaccines, is hampered by underreporting, over reporting and reporting bias. This makes it difficult to identify the true incidence of these events and the presence of multiple confounders which may not enable the assessment of the causality with higher specificity.^22^ In addition, due to the nature of the database, it was not possible to know the denominator (the total number of vaccinated individuals for each type of vaccine), which hinders the analysis of likelihood of true occurrence of thrombotic adverse events for each vaccine. Also, we were unable to report possible concomitant drugs. Despite these limitations, the study used global “real-world” data and collected valuable information about three widely used vaccines, where more than two thirds of all reports were received from healthcare professionals which increases the credibility and quality of reports.

People who are vaccine hesitant and reluctant to take any of the mentioned vaccines,^23^ should know that most COVID-19 vaccines are effective to prevent symptomatic infection including hospital admissions and severe disease.^24,25^ The risk of COVID-19 related thrombotic events are minimal and likely manageable with available treatments.^8^ Thrombotic adverse events reported for the three vaccines remains extremely rare.^8,26^

In summary, thrombotic adverse events reported for the three vaccines remains extremely rare with multiple causative factors reported elsewhere as precipitating these events. Practicing vigilance and proper clinical management for the affected vaccines, as well as continuing to report adverse events, are essential.

## Data Availability

The dataset is in the public domain

## Data sharing

All data are available from the public domain portal at adrreports.eu

## Contributors

Conceptualisation MT and HE; formal analysis HE; methodology MT and HE, project administration MT and HE; validation KM and DS; writing – original draft MT and HE; writing – substantial review, rewriting & editing KM, MT and DS.

